# Machine Learning Model for Predicting Number of COVID19 Cases in Countries with Low Number of Tests

**DOI:** 10.1101/2021.07.12.21260298

**Authors:** Samy Hashim, Sally Farooq, Eleni Syriopoulos, Kai de la Lande Cremer, Alexander Vogt, Nol de Jong, Victor L. Aguado, Mihai Popescu, Ashraf K. Mohamed, Muhamed Amin

## Abstract

The COVID-19 pandemic has presented a series of new challenges to governments and health care systems. Testing is one important method for monitoring and therefore controlling the spread of COVID-19. Yet with a serious discrepancy in the resources available between rich and poor countries not every country is able to employ widespread testing. Here we developed machine learning models for predicting the number of COVID-19 cases in a country based on multilinear regression and neural networks models. The models are trained on data from US states and tested against the reported infections in the European countries. The model is based on four features: Number of tests Population Percentage Urban Population and Gini index. The population and number of tests have the strongest correlation with the number of infections. The model was then tested on data from European countries for which the correlation coefficient between the actual and predicted cases R^2^ was found to be 0.88 in the multi linear regression and 0.91 for the neural network model. The model predicts that the actual number of infections in countries where the number of tests is less than 10% of their populations is at least 26 times greater than the reported numbers.

## Introduction

The SARS-CoV-2 or COVID-19 outbreak was declared a global health emergency on the 30th of January 2020 by the WHO. COVID-19 is a member of the coronavirus family enveloped positive sense single stranded RNA viruses. It is thought COVID-19 made the transition from animal to human hosts on the Huanan seafood market in Wuhan in the province of Hubei China.^1^ The virus spread rapidly initially within China and then Worldwide. COVID-19 was declared a pandemic on the 11th of March 2020 by the World Health Organization. As of April 25th, 2021 there have been almost 100 million confirmed cases worldwide. Yet PCR (polymerase chain reaction) which can detect the genetic material of the virus is the most accurate technique for identifying the COVID19 infections.^2^

COVID-19 has exposed several inequalities. In the scrabble to obtain medical resources poorer countries have been left behind. Governments of low and middle income countries have struggled to provide sufficient funds to obtain medical resources such as COVID-19 tests.^3^ Furthermore more geo-politically powerful countries have been accused of hoarding supplies leaving poorer countries unable to access sufficient tests.^4^ With a disparity in the number of COVID-19 tests available we aim to provide a prediction model based on machine learning that mitigates the reliance on clinical tests.

Machine learning has been utilized in contact tracing as a diagnostic and prognostic tool in vaccine and treatment development as a method to forecast and predict COVID-19 cases and deaths.^5-11^ It has the potential to reduce the strain on healthcare systems that have been heavily burdened by the COVID-19 Pandemic. For example machine learning has been used to predict a positive COVID-19 infection in a PCR test.^12^ The prediction is based on 8 binary features including age sex contact with individuals known to have had COVID-19 and including the appearance of five clinical symptoms. In addition, Sun et. al., developed a model to predict the severity of a COVID-19 infection.^13^ Furthermore a model is utilized to predict the number of COVID-19 patients between one and six days in advance in 10 Brazilian states.^14^

In this work we build a multilinear regression and a neural network models to predict the number of COVID19 as of 15/03/2021. The models are trained on the US states data and tested against the number of infections in the European countries. Then both were used to predict the COVID-19 infections in countries with low number of tests. The model is based on four features: the number of tests population urban population and the Gini index. The model suggests that the actual number of infections is at least 10 times higher than the reported numbers of infections.

## Results and discussion

Since the start of the COVID-19 pandemic the US has conducted over 400 million COVID-19 tests making the country a rich and reliable source of information.^15^ For this reason the data from all US states was used to train our machine learning models. To evaluate the models, they were tested against the data from the European countries. Finally, the models are used to make predictions for the number of COVID-19 cases in countries that have conducted low numbers of tests. The following countries are used as an example for low-testing countries: Nepal Vietnam Mongolia Kenya Ghana Zambia Iran Paraguay and Ecuador.

### Features Analysis

The features currently utilized in the models are: ‘Population’ ‘Tests’ ‘Gini’ and ‘% urban population’. To observe their collinearity, the number of cases were plotted against these features for the US states (Figure 1).

**Figure 1.**
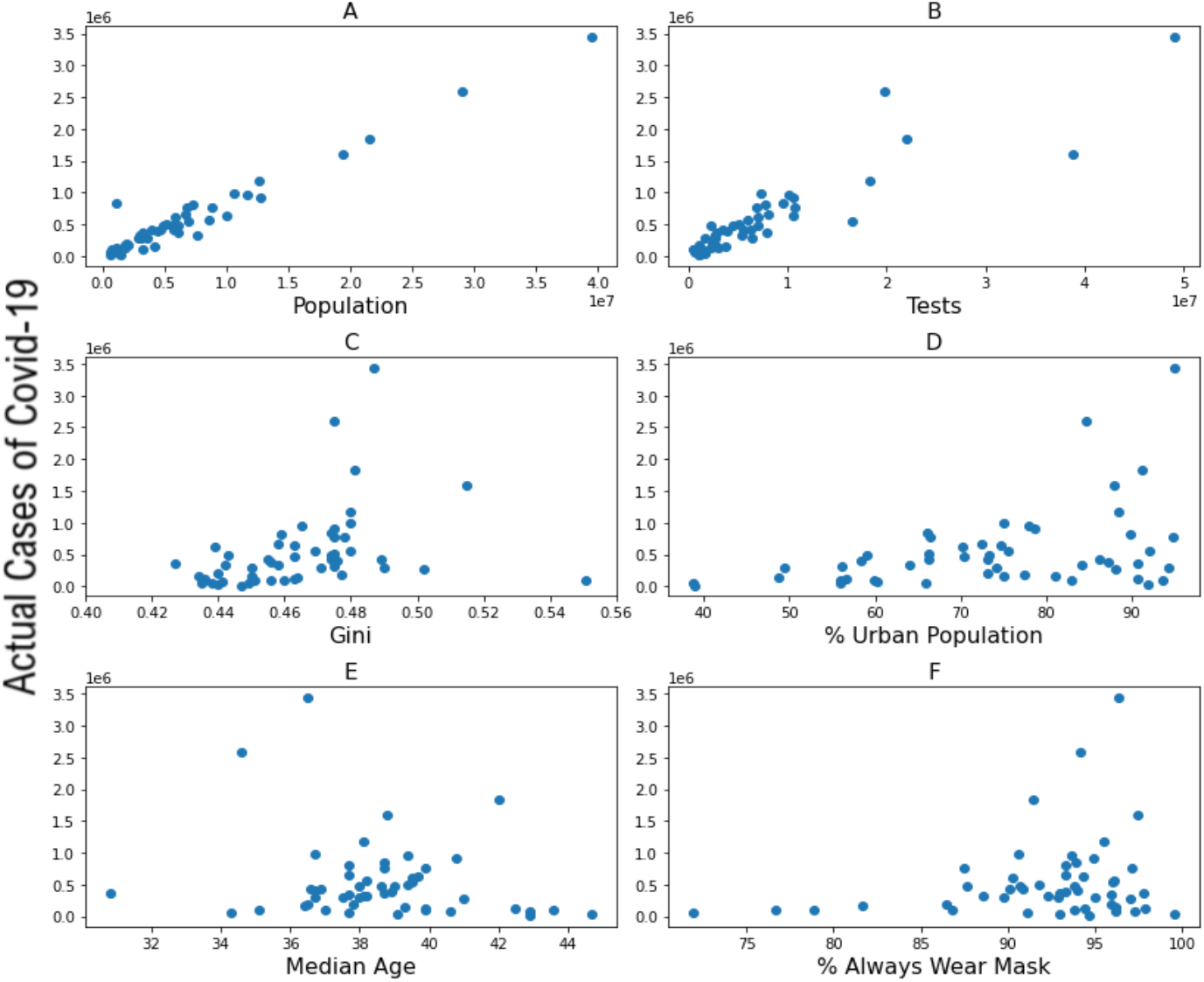
COVID-19 Cases vs. A) Population. B) Number of tests. C) Gini. D) % Urban Population E) Median Age F) % of Population that always wears a mask. Each point represents a state.

The population and the number of tests conducted both show strong correlation with the number of COVID-19 cases with R^2^ values of 0.95 and 0.81 respectively (Figure 1 (AB)) and p values of zero. However, a much lower correlation was obtained for the Gini index and percentage urban population with R^2^ values of 0.12 and 0.16 and p values of 0.01 and 0.003 respectively. The features that are currently utilized in the models were selected based on their strong correlation with the number of cases. Other features such as ‘Median Age’ ‘% of people wearing a facemask outside’ ‘Number of lockdown days’ were not used as low correlation was found between these features and number of cases and because the data was incomplete for a number of these features. Adding these features to the models would have resulted in a higher error.

### Multilinear regression

A multiple linear regression model was built and trained on the US states data according to the following equation:

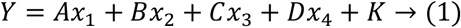

### Multilinear regression

Where *Y* denotes the number of cases A B C and D are the regression coefficients obtained from least square fitting *x*_1_ *x*_2_ *x*_3_ *and x*_4_ are the independent variables (populations number of tests Gini index and % of urban population respectively) and K is the y-intercept.

The model shows a very strong correlation between the predicted and actual number of COVID-19 cases for both the US states data (the training dataset) and the European data (test dataset) (Figure 2).

**Figure 2.**
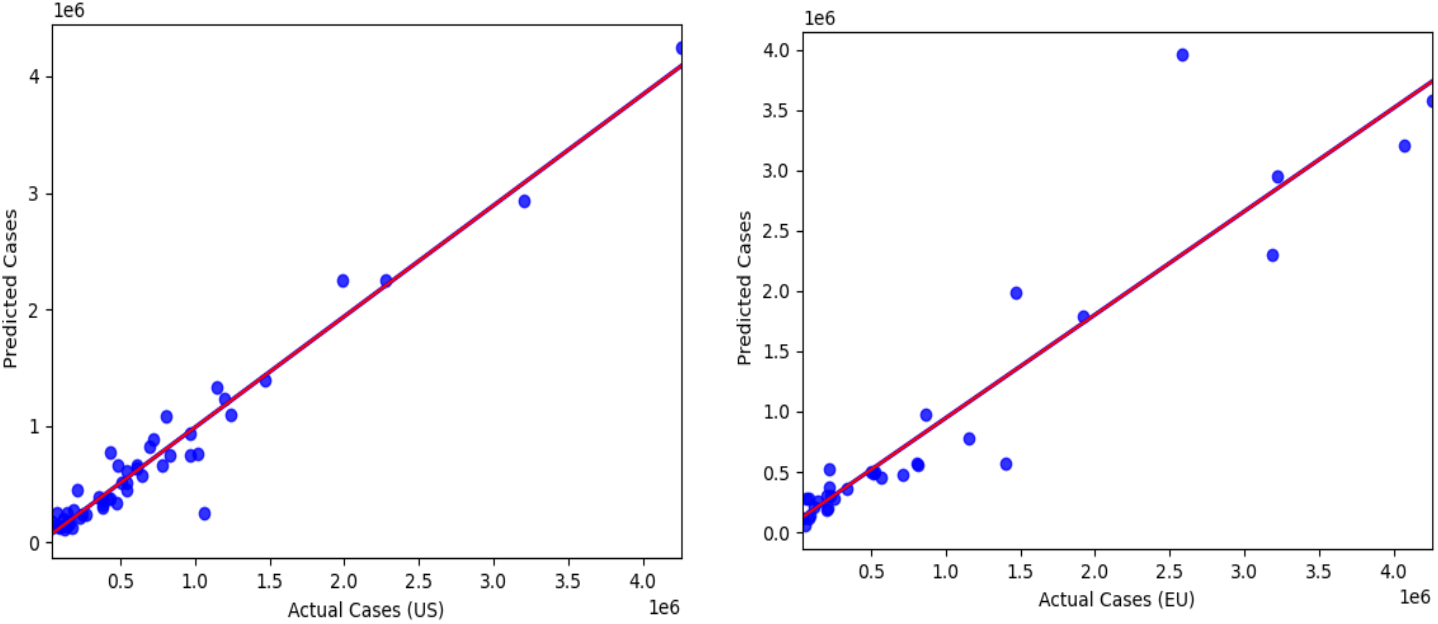
To the left the predictions vs observed cases for US data (Slope: 1.00 Intercept: 0 R^2^: 0.95). To the right the predictions vs observed cases for European data (Slope: 1.49 Intercept: 12K R^2^: 0.88)

For the US data the calculated slope is 1.00 with an intercept of zero and R^2^ of 0.95. For the European data the correlation coefficient R^2^ is 0.88 and the slope and the intercepts are 1.49 and 12k respectively, which indicates that the predicted number of infections for the EU is generally higher than the reported. This could result from the differences in the behavior and commitment of the people toward the governmental rules in the US and the EU.

To understand the contribution of each feature to the prediction model we report the estimated regression coefficients for each of the four features. The calculated coefficients are 0.87 0.13 -0.01 and -0.03 for the populations number of tests Gini and % urban population respectively.

The ‘population’ feature has a score close to one and thus is the major contribution to the prediction model. The scores for the ‘% Urban Population’ and ‘Gini’ are negative which suggests that these features are not significant for the regression model.

### Neural networks

The neural network model is mainly considered to account for possible nonlinearities in the Gini index and percentage of urban populations. A fully connected Deep Neural Network (DNN) is trained and tested with US and EU datasets respectively. The input layer of the network consists of 128 nodes and is followed by four hidden layers with 128 nodes and an output layer with a single node. The number of nodes of the output layer corresponds to the number of classes. Each layer has a random weight and bias initialization based on the normal distribution initializer which is necessary to set the first set of numbers of weights and biases and thus kick off the training procedure. The ReLU function has become the default activation function for many types of neural networks because such models are easy to train and often achieve good performance.

The DNN model is trained with an objective function (loss function) which needs to be minimized. The Mean Squared Error (MSE) is used as a loss function and Stochastic Gradient Descent (SGD) optimizer is employed to find the best values for the DNN parameters by minimizing the loss function iteratively over the dataset. The number of iterations (epochs) is chosen to be 100 epochs. The network is trained using data from US states and tested using data from European countries using the same set of features as in the case of multilinear regression namely Population Tests Gini and the percentage of urban population. The testing results is illustrated in (Figure 3) which quantifies the correlation between the predicted number of infections and the number of infections recorded. The slopes are 0.95 and 0.80, the values are 0.95 and 0.91 and the mean absolute error of 0.03 and 0.06 for the US and EU datasets respectively. These measurements suggest that the model fits the observed data by learning the relationships between the input variables.

**Figure 3.**
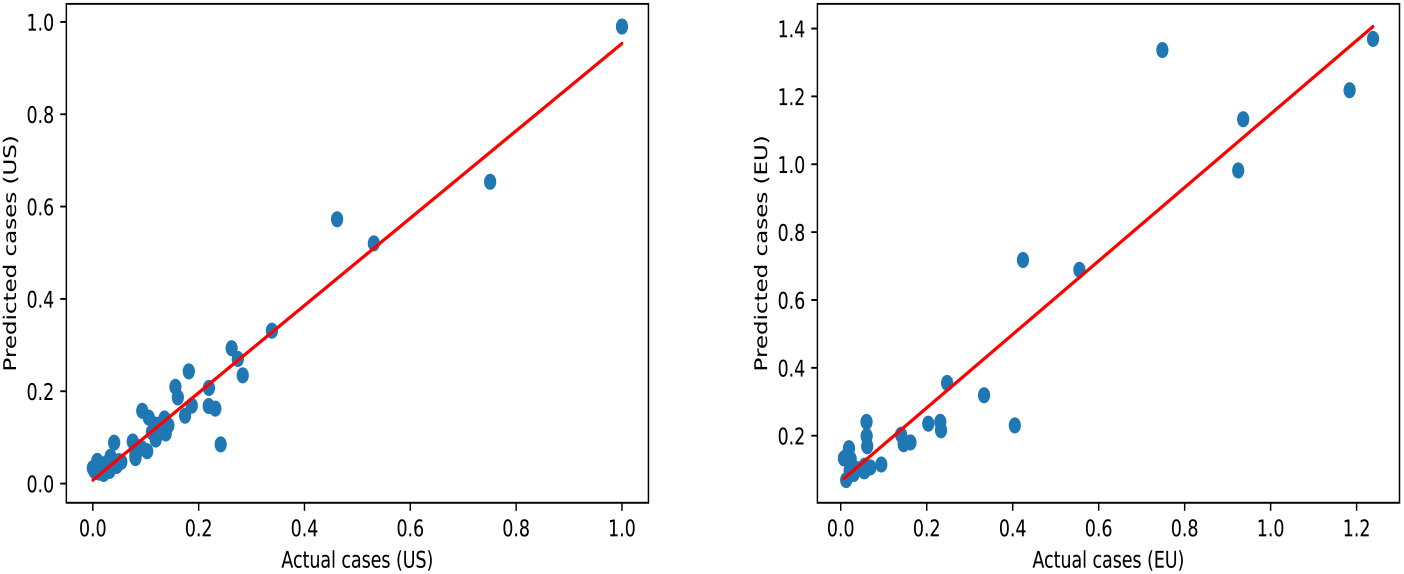
A) the predictions vs observed cases for US data (Slope: 0.95 Intercept: 0.0 R^2^: 0.95) B) the predictions vs observed cases for European data (Slope: 1.57 Intercept: 45K R^2^: 0.81)

### Prediction of COVID-19 cases

The reported infections and their corresponding predicted values (using linear regression and NN) are shown in table 1. Furthermore, according to the training dataset the US has performed 361 million tests which is equal to approximately 110% of the US population. Thus, we reported the predicted number of cases for European and other countries with low number of tests as if these countries have had tests equal to 1.1 multiplied by their respective populations (columns 6-8 of table 1). Although the number of tests for the EU countries is increased by 30%, the slopes of the linear regression and the NN models are increased only by 5% and 11% respectively.

**Table 1.**
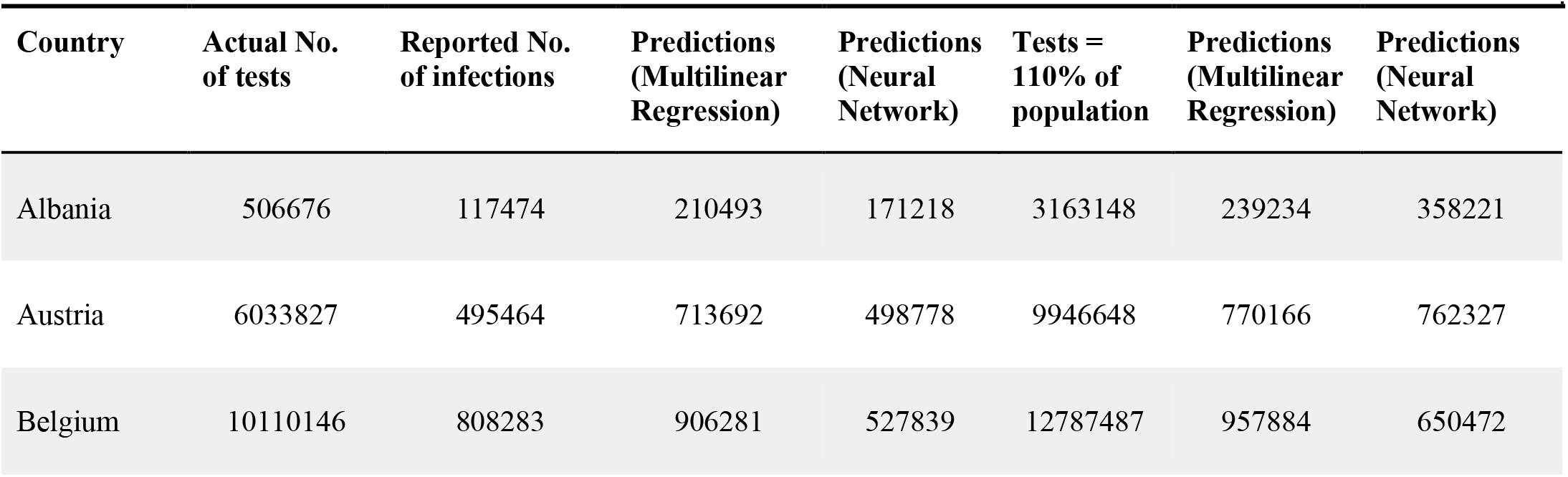
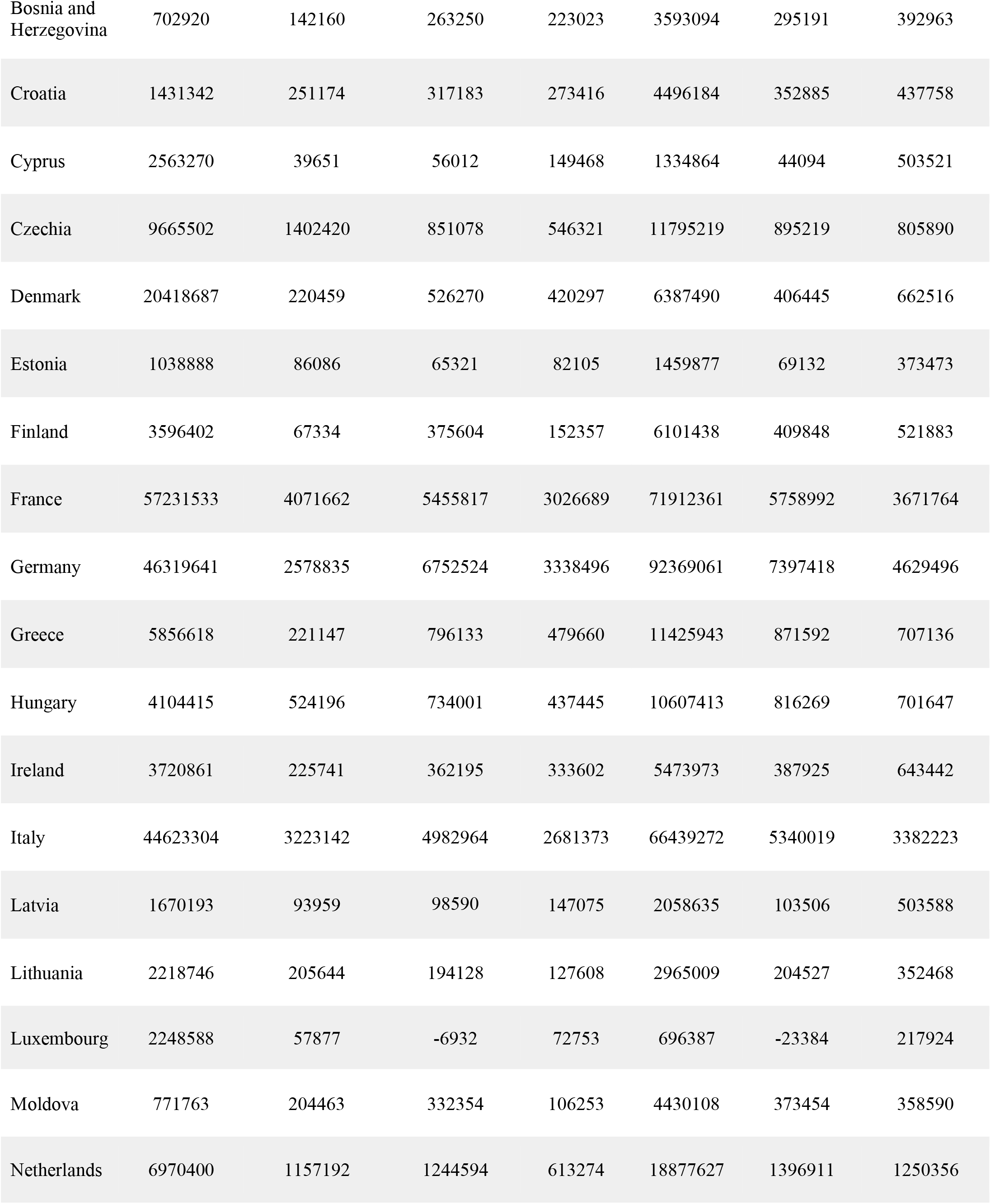
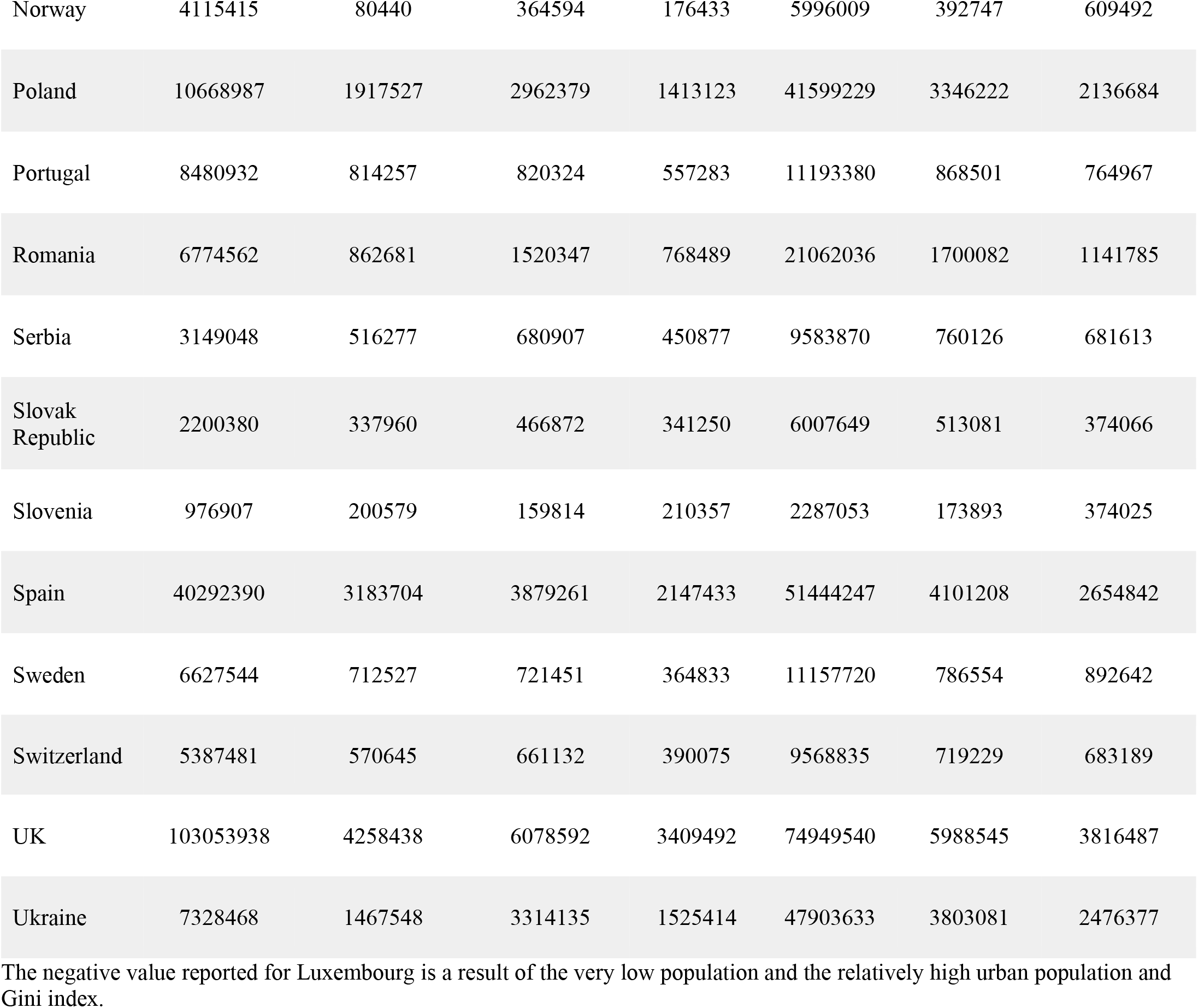
The predicted number of COVID-19 cases for test countries

| Country | Actual No. of tests | Reported No. of infections | Predictions (Multilinear Regression) | Predictions (Neural Network) | Tests = 110% of population | Predictions (Multilinear Regression) | Predictions (Neural Network) |
| --- | --- | --- | --- | --- | --- | --- | --- |
| Albania | 506676 | 117474 | 210493 | 171218 | 3163148 | 239234 | 358221 |
| Austria | 6033827 | 495464 | 713692 | 498778 | 9946648 | 770166 | 762327 |
| Belgium | 10110146 | 808283 | 906281 | 527839 | 12787487 | 957884 | 650472 |
| Bosnia and Herzegovina | 702920 | 142160 | 263250 | 223023 | 3593094 | 295191 | 392963 |
| Croatia | 1431342 | 251174 | 317183 | 273416 | 4496184 | 352885 | 437758 |
| Cyprus | 2563270 | 39651 | 56012 | 149468 | 1334864 | 44094 | 503521 |
| Czechia | 9665502 | 1402420 | 851078 | 546321 | 11795219 | 895219 | 805890 |
| Denmark | 20418687 | 220459 | 526270 | 420297 | 6387490 | 406445 | 662516 |
| Estonia | 1038888 | 86086 | 65321 | 82105 | 1459877 | 69132 | 373473 |
| Finland | 3596402 | 67334 | 375604 | 152357 | 6101438 | 409848 | 521883 |
| France | 57231533 | 4071662 | 5455817 | 3026689 | 71912361 | 5758992 | 3671764 |
| Germany | 46319641 | 2578835 | 6752524 | 3338496 | 92369061 | 7397418 | 4629496 |
| Greece | 5856618 | 221147 | 796133 | 479660 | 11425943 | 871592 | 707136 |
| Hungary | 4104415 | 524196 | 734001 | 437445 | 10607413 | 816269 | 701647 |
| Ireland | 3720861 | 225741 | 362195 | 333602 | 5473973 | 387925 | 643442 |
| Italy | 44623304 | 3223142 | 4982964 | 2681373 | 66439272 | 5340019 | 3382223 |
| Latvia | 1670193 | 93959 | 98590 | 147075 | 2058635 | 103506 | 503588 |
| Lithuania | 2218746 | 205644 | 194128 | 127608 | 2965009 | 204527 | 352468 |
| Luxembourg | 2248588 | 57877 | -6932 | 72753 | 696387 | -23384 | 217924 |
| Moldova | 771763 | 204463 | 332354 | 106253 | 4430108 | 373454 | 358590 |
| Netherlands | 6970400 | 1157192 | 1244594 | 613274 | 18877627 | 1396911 | 1250356 |
| Norway | 4115415 | 80440 | 364594 | 176433 | 5996009 | 392747 | 609492 |
| Poland | 10668987 | 1917527 | 2962379 | 1413123 | 41599229 | 3346222 | 2136684 |
| Portugal | 8480932 | 814257 | 820324 | 557283 | 11193380 | 868501 | 764967 |
| Romania | 6774562 | 862681 | 1520347 | 768489 | 21062036 | 1700082 | 1141785 |
| Serbia | 3149048 | 516277 | 680907 | 450877 | 9583870 | 760126 | 681613 |
| Slovak Republic | 2200380 | 337960 | 466872 | 341250 | 6007649 | 513081 | 374066 |
| Slovenia | 976907 | 200579 | 159814 | 210357 | 2287053 | 173893 | 374025 |
| Spain | 40292390 | 3183704 | 3879261 | 2147433 | 51444247 | 4101208 | 2654842 |
| Sweden | 6627544 | 712527 | 721451 | 364833 | 11157720 | 786554 | 892642 |
| Switzerland | 5387481 | 570645 | 661132 | 390075 | 9568835 | 719229 | 683189 |
| UK | 103053938 | 4258438 | 6078592 | 3409492 | 74949540 | 5988545 | 3816487 |
| Ukraine | 7328468 | 1467548 | 3314135 | 1525414 | 47903633 | 3803081 | 2476377 |

The negative value reported for Luxembourg is a result of the very low population and the relatively high urban population and Gini index.

Using the same training dataset, we predicted the number of infections in selected countries where the number of tests is less than 10% of their populations (table 2). The average number of the predicted infections is higher than the reported by 26 times for the linear regression model and 4 times for the NN. The discrepancy between the results from multilinear regression and NN models in table 2 is due to the overfitting feature of the NN. The overfitting indicates that the generalization of the NN model is rather limited. This is due to the minimal dataset, 52 entries, used for the training procedure, which is not enough for the NN model to avoid overfitting.

**Table 2.** The predicted number of COVID-19 cases for countries whose total tests equal less than 10% of their population

| Country | Actual No. of tests | Reported No. of infections | Predictions (Multilinear Regression) | Predictions (Neural Network) | Tests = 110% of population | Predictions (Multilinear Regression) | Predictions (Neural Network) |
| --- | --- | --- | --- | --- | --- | --- | --- |
| Afghanistan | 465731 | 55985 | 3034780 | 404687 | 41845929 | 3422007 | 509312 |
| Algeria | 230861 | 115410 | 3327210 | 349522 | 47358359 | 3768219 | 470700 |
| Chad | 119517 | 4328 | 1317840 | 158913 | 17541564 | 1480872 | 219913 |
| DRC | 159469 | 27077 | 6681106 | 779441 | 95469624 | 7572999 | 1110283 |
| Egypt | 2824316 | 191555 | 7770111 | 1100450 | 110426880 | 8777033 | 1318575 |
| Guatemala | 1411568 | 183014 | 1316936 | 194200 | 18264429 | 1474642 | 164909 |
| Honduras | 714929 | 178925 | 776194 | 98998 | 10720729 | 869826 | 85309 |
| Indonesia | 16610468 | 1430000 | 20837478 | 3404783 | 297688125 | 23467745 | 3339198 |
| Mozambique | 454528 | 64516 | 2372216 | 270953 | 33402640 | 2680538 | 387939 |
| Pakistan | 9530000 | 609964 | 16699868 | 2557415 | 238221850 | 18839920 | 2686825 |
| Papua New Guinea | 112995 | 2269 | 792772 | 127738 | 9653720 | 882052 | 176443 |
| Syria | 103566 | 16556 | 1357616 | 136954 | 18777149 | 1532360 | 158446 |
| Yemen | 62990 | 2908 | 2313264 | 271150 | 32078114 | 2612855 | 369019 |

All the numbers are reported up to 15/03/2021
\* Where no test data could be found for 15/03/2021 data up till 31/05/2021 was used

## Conclusions

Both the multilinear regression and neural network models predicted the number of COVID-19 cases with a fair degree of accuracy on the European test data set. Considering table 1 the number of cases predicted by the models were close to the number of cases reported for some countries for Italy Poland and Slovakia. Yet in most cases the model predicted more cases than were reported. The models were trained on data from the US a country that tested extensively. Therefore, it seems that due to limited testing in most countries the number of cases reported are a gross underestimation of actual number of infections. This disparity was most pronounced in countries that are not testing extensively. The predicted number of infections for these countries is 26 times higher than the reported numbers on average. Therefore, the models can be effective tools for estimating the number of COVID-19 infections in countries where sufficient testing is not available or where it is suspected that governments may not be being entirely transparent about the number of COVID-19 infections.

## Methods

The data was obtained from several official sources for example from the World Bank World Development Indicators^16-19^ government websites and publications^20-22^ Worldometer^23^ and from Our World in Data^14^. This data was extracted standardized and compiled into a single file. Although several features were considered only four were included in the model owing to a lack of availability of data and low correlation with COVID-19 cases recorded. The four features used were: Population Tests Gini Index % Urban Population. As the model first needed to be trained on US states and then tested on European countries data for all factors included would need to be available for both. This considerably limited the number of features that could be incorporated into the models. Several other factors were also considered for example median age and percentage of the population that always wears a face mask. However median age was excluded from the model as it had poor correlation with the number of infections. The mask wearing variable was excluded as the proportion of the populations that always wore masks was measured differently between the training and test countries and likely with all other countries for which the models were used to make predictions.

Data used to train the model covered the period from the beginning of the pandemic to February 2021. Later data was not used owing to the vast differences among countries not only in the starting date and accessibility of vaccines but also the rate of vaccination. These discrepancies would make predictions for other countries inaccurate. The data used to test the model covered the period up until March 15^th^, 2021. A later date was considered for the test data than for the training data as most European countries started vaccination after the US.

Although the intention was originally to train the data on Indian states as well as US states to allow for different models for the developing and developed countries. India was excluded owing to the high prevalence of the new B. 1.617 variant which has increased transmissibility^24^. Although replacing India with Russia as an additional training data set was considered the lack of data available made this unfeasible.

Some pre-processing steps had to be taken to clean the data before it could be used for the machine learning algorithm. First, the relevant features and information were extracted from the .csv file where the data is stored whereupon all commas were removed from individual data points to make sure python could parse them correctly. The data was then normalized via a min-max-scaler which places all data points between 0 and 1. For each data point in a feature the MinMaxScaler deducts the smallest value in the feature and then divides this answer by the range which is the difference between the original maximum and original minimum. The MinMaxScaler retains the original shape of the distribution thus preserving the information embedded into the initial data set. However, it is important to note that this also means that the MinMaxScaler does not reduce the importance of outliers. Finally, the pre-processing procedure was completed by removing data samples that had missing values for some of their features. This is to make sure that all data can be used for training the model as missing values can cause errors and unwanted variations within the procedure.

Two different types of machine learning algorithms were used for analysis on the data multi-linear regression and a multi-layer perceptron artificial neural network (ANN). The multiple linear regression model was built using Scikit-learn library^16^. The neural network code operates Keras architecture from the Tensorflow^25^ library to construct the model. The ANN utilizes 1 output layer 1 input layer and 3 dense hidden layers visualized in the following figure:

**Figure 7.**
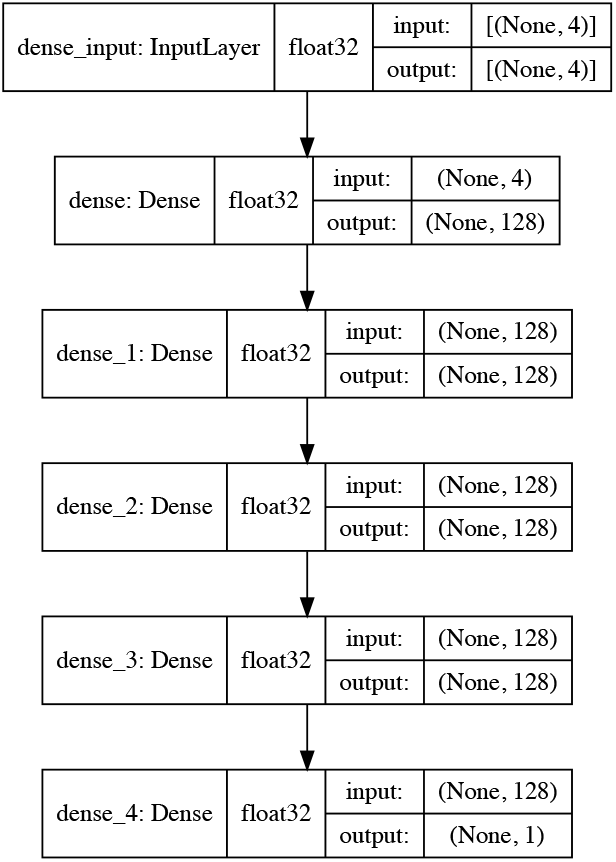
Artificial Neural Network Architecture

All dense layers are using the Rectified Linear Unit (ReLU) as activation function which is defined as follows:

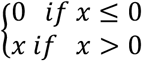

The slope is always 0 for negative inputs and always 1 for positive inputs. ReLU was used as it is computationally less intensive and faster than most other activation functions such as sigmoid and tanh.

The mean squared error (MSE) function is used to calculate loss in the current iteration of the neural network. This function takes the absolute error of all points and calculates their mean. MAE is calculated via the following equation:

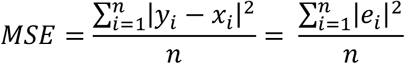

MSE was used because it is a commonly used metric and relatively robust to outliers which suitable for the data used in this study.

The neural network contains a few hyperparameters that had to be set manually before the training. These hyperparameters are chosen by using a random grid search technique. The choice of the ReLU activation function the number of hidden layers and the number of nodes in each layer are examples of hyperparameters.

## Data Availability

All the data are available on the referenced websites

